# Primary healthcare capacity in Northwest Ethiopia: Insights through the Primary health care progression model

**DOI:** 10.1101/2024.12.22.24319511

**Authors:** Chalie Tadie Tsehay, Nigusu Worku, Endalkachew Dellie, Wubshet Debebe Negash, Andualem Yalew Aschalew, Ayal Debie, Tsegaye G. Haile, Samrawit Mihret Fetene, Adane Kebede, Asmamaw Atnafu

## Abstract

**Background:** Primary healthcare (PHC) systems are widely recognized as essential foundations for ensuring equitable access to quality medical care for all. Achieving the health-related Sustainable Development Goals (SDGs), including the sub-targets of universal health coverage by 2030 requires resilient PHC systems, supported by scientific evidences to inform better policy. However, there is a lack of evidence regarding the PHC system capacity at the operational level in Ethiopia. Therefore, we assessed the capacity of primary health care at the health facilities level in northwest Ethiopia.

**Methods:** We used a mixed-method assessment of the PHC capacity guided by the progression model, which includes governance, input, and population health and facility management domains with a total of 33 rubric-based (scaled from 1 to 4) measurement items. We included a total of three primary hospitals and five health centers from Northwest Ethiopia. Key informants interviews, facility observations including guideline and policy reviews and reports, discussion with key stakeholders, were our source of data. Data were independently collected by two groups of assessors (internal and external assessors) and a final score was determined by consensus through panel discussion. Finally, we summarized and synthesized the results over the three domains of PHC capacity assessment and the nine subdomains.

**Results:** All the three domains scores were found to be low. We found that the scores were 1.5, 2.2, and 1.3 out of four points for the governance, input, and population health and facility management domains, respectively. While we found a better achievement on health management information system and civil registration and vital statistics, the local priority setting, facility management capability, innovation and learning, community engagement and social accountability measures had lowest capacity score.

**Conclusions:** Our study highlighted that the governance and population health and facility management domains scored lower at the health facilities in central Gondar zone. Therefore, it is crucial to enhance these domains to strengthen PHC though a comprehensive approach, aiming to meet its targets and achieve UHC by 2030 or beyond.

## Background

Primary healthcare (PHC) system is the foundations for guaranteeing that everyone has equitable access to quality medical care[1]. Sustaining a strong PHC system is also crucial for meeting the health-related Sustainable Development Goals (SDG) and its sub-targets of universal health coverage (UHC) by 2030 or beyond[2].

The Astana Declaration of 2018 emphasized the critical role that PHC must play in achieving UHC and reiterated the international community’s commitment to developing PHC[3]. But all too often, especially in low- and middle-income countries (LMICs), PHC is ineffective, under prioritized, and unable to fulfill its promise[4].

The PHC system, which emphasizes service accessibility by delivering it as near to people’s homes and communities as possible, acts as the cornerstone of the health system[5]. Accordingly, Ethiopia has taken a number of steps to address the fragmentation of basic health cares through a community-based PHC services.

Despite a wide range of interventions aimed at improving PHC capacity, there is limited evidence regarding the impact of primary healthcare system performance across country [6]. The Primary Health Care Performance Initiative (PHCPI) was developed to measure PHC and evaluate PHC systems[7]. Owing to the lack of available data sources and metrics for PHC capacity, PHCPI created the PHC Progression Model, a participatory rubric-based mixed-methods evaluation tool, to measure capacity [8].

The lack of resources and practical implementation strategies has created a need to accelerate progress in achieving universal health goals[9]. The health system delivers preventive, promotive, curative, and rehabilitative interventions through a combination of public health actions and a pyramid of healthcare facilities that provide personal healthcare[10]. The PHC facilities play a crucial role in addressing essential health care. Despite limited national-level studies, there remains a lack of evidence regarding the capacity of PHC at the zonal and district level. Therefore, we evaluated the capacity of primary health care at the health facility level in Northwest Ethiopia.

## Methods

### Study context

Our study was conducted from March to June 2023 in Central Gondar zone of northwest Ethiopia, which comprises one comprehensive specialized hospital, one general hospital, nine primary hospitals, 76 health centers and 154 health posts. The study focused on three randomly chosen public primary hospitals (Wogera, Sanja, and Qoladba) and five health centers (Ambagiorgis, Makisegnit, Qoladba, Chua hit, and Sanja).

### PHC capacity assessment

In our study, to evaluate the PHC capacity, we used the PHC progression model taken from PHC performance initiative[11]. This progression model was recently introduced to systematically guide PHC capacity assessment, with the aim of improving PHC system performance in resource-limited settings and contributing to the achievement of UHC targets in the long run.

The capacity aspect of the PHC-VSP evaluates three main elements of primary healthcare: governance, inputs, and management of population health and facilities. The assessment tool for capacity assessment has 33 specific measures derived from the primary healthcare progression model[7].

### Data sources and assessment procedures

We used multiple data collection methods from different data sources including key informant interviews, documents (patient charts and registration) reviews, and facility observations (review of facilities’ strategic plan, reports). We adapted tools for secondary data collection regarding to surveillance and facility inputs from the Ethiopian Public Health Institute’s (EPHI) Service Availability and Readiness Assessment (SARA) [12] In addition, we utilized the DHIS 2 databases, from hospital and health center levels to address matters related to population health and health facility management.

The evaluation process first encompassed an internal review conducted by healthcare facility staff, followed by external assessments carried out by evaluation teams from outside the study areas. The 33 individual measure scores are consolidated into nine sub-scores, each representing a subdomain of the PHC Capacity. These sub-scores are calculated by taking the simple unweight average of the relevant measures. The nine sub-scores are further aggregated into three overall scores—Governance, Inputs, and Population Health and Facility Management—which are displayed in the Capacity pillar of the Vital Signs Profile (Table 1). These overall scores are also calculated using a simple unweight average of the respective sub-scores.

**Table 1:** The primary health care progression model vital signs in three main domains, central Gondar, Ethiopia, 2023.

| VSP Capacity Score | Category | Score |
| --- | --- | --- |
| **Governance** | OVERALL GOVERNANCE | 1.5 |
|  | Governance and Leadership | 1.6 |
|  | Adjustment to Population Health Needs | 1.3 |
| **Inputs** | OVERALL INPUTS | 2.2 |
|  | Drugs and Supplies | 2.0 |
|  | Facility Infrastructure | 2.3 |
|  | Information Systems | 2.7 |
|  | Workforce | 2.5 |
|  | Funds | 1.7 |
| **Population Health and Facility Management** | OVERALL POPULATION HEALTH AND FACILITY MANAGEMENT | 1.3 |
|  | Population Health Management | 1.0 |
|  | Facility Organization and Management | 1.6 |

164 Color code: Yellow=2-3 and pink = less than 2

#### Document review and synthesis

A PHC capacity assessment team, consisting of two members assigned to each of the eight health facilities. Utilizing different templates specifically developed for the assessment, the team gathered relevant data from different sources.

#### Key informant interviews

After review of documents and finalizing the synthesis of secondary data, the assessment team thoroughly identified information gaps and indicators requiring additional verification and clarification subsequently, the team identified key informants based on their exposure and expertise.

### Internal assessment

The internal assessors were recruited with in each facility considering their experience. Two days of training was given for internal assessors on data collection tools, study objective, participant handling and other ethical issues, and they have measured the capacity of the PHCs from their respective health facilities using both document review and key informants interview and gave scores for all 33 measures.

### External Assessment

We recruited a total of two external assessors, all of whom held master’s degrees in public health. The recruitment process was considered the assessors previous data collection and related experience. Prior to the actual data collection, two days training was given for data collectors. Like internal assessors the team gathered pertinent data from their assigned health facilities, and the collected data were combined using a template developed for this purpose. The scoring scale was the same for internal and external assessors (score of 1 to 4 for each measure).

### Data synthesis

The external assessment team combined relevant information derived from document review, desk review and key informant interviews related to the 33 measures.

This information was briefly summarized and organized into a single document for use in both internal and external assessments. The internal and external assessment teams evaluated the 33 PHC capacity measures based on this evidence summary, recording the scores in an Excel file. The findings taken from each data source were documented separately, with clear indications of their sources. Both qualitative and quantitative data from various sources were recorded and combined for each of the 33 measures, using data synthesis template for respective facilities. Scoring in the PHC Progression Model follows a threshold approach, where a performance level is only achieved if all criteria within a measure meet the standards outlined in the corresponding rubric. Each indicator was given rating scale of one to four, based on criteria’s taken from the PHC progression model. During rating scale the lowest capacity was considered as one whereas the highest capacity score was scored as four. Subsequently, these scores were aggregated into nine sub-domains and three domains of PHC capacity assessment. In this study, we figure out the summarized findings for each domain and sub-domain of the assessment. The key informant interviews were recorded, then coded, transcribed and translated to be suitable for analysis.

### Ethical considerations and consent to participate

Ethical review, approval and clearance were granted by the Institutional Review Board (IRB) of the University of Gondar (Ref.no R/T/T/T/C/ENG./193 /11/2022) and an official letter of support was obtained from Gondar city administration Health Department. Permission letter was also taken from each health facility. All methods were performed in accordance with the declaration of Helsinki. Written informed consent to participate was taken from each study participant. Thereby each respondent signed consent after a brief explanation of the risk and benefits of their involvement in the study. Participants were also informed about their right to withdraw from the study at any time they want and/or to skip questions that they are not comfortable to respond.

## Results

### Primary health care progression model vital signs (VSP)

The assessment of PHC capacity in central Gondar health facilities revealed that all most below the expected standard. In the governance domain, the average score is 1.5, out of 4. Regarding the inputs average score domain, 2.2. On the other hand, in the population health and facility management domain, the PHC capacity scored is 1.3 (Table 1).

### Governance

Four measures namely primary healthcare policy, primary healthcare leadership, social accountability and multi-sectorial actions were assessed under Governance domain. The assessment findings showed that Primary health care policies had medium score (3/4) whereas the rest three measures had low score (1/4) each.

### Inputs

Most of the indicators to assess the input domain were taken from Service Availability and Readiness Assessment (SARA) report since 2028. Accordingly, five main domains were selected for this study. As the finding indicated the fund domain inputs had lost score (1.7/4) whereas the rest four had medium score (2-2.7/4).

### Population Health and Management

The characteristics of Ethiopia’s health system within the population health and facility management domain are marked by features such as community engagement in health planning, implementation and evaluation. Community engagement is raised through different initiation like Women’s health Development Armies, Managing board at health institution as well as community-based health insurance. Two main domains (population health management and facility management) were assessed under population health and facility management. The result showed that both domains had low scores (1 and 1.6/4) respectively.

### Comparison of Capacity assessment scores across health facilities

Among the 33 measures in the Primary Health Care Progression Model, only the health information management measure achieved a higher score (3 out of 5 for some health facilities and 3 out of 4 for others), indicating a yellow or green code. However, three measures, namely social accountability, innovation and learning, and empanelment, had the lowest scores across all facilities (Table 2).

**Table 2:**
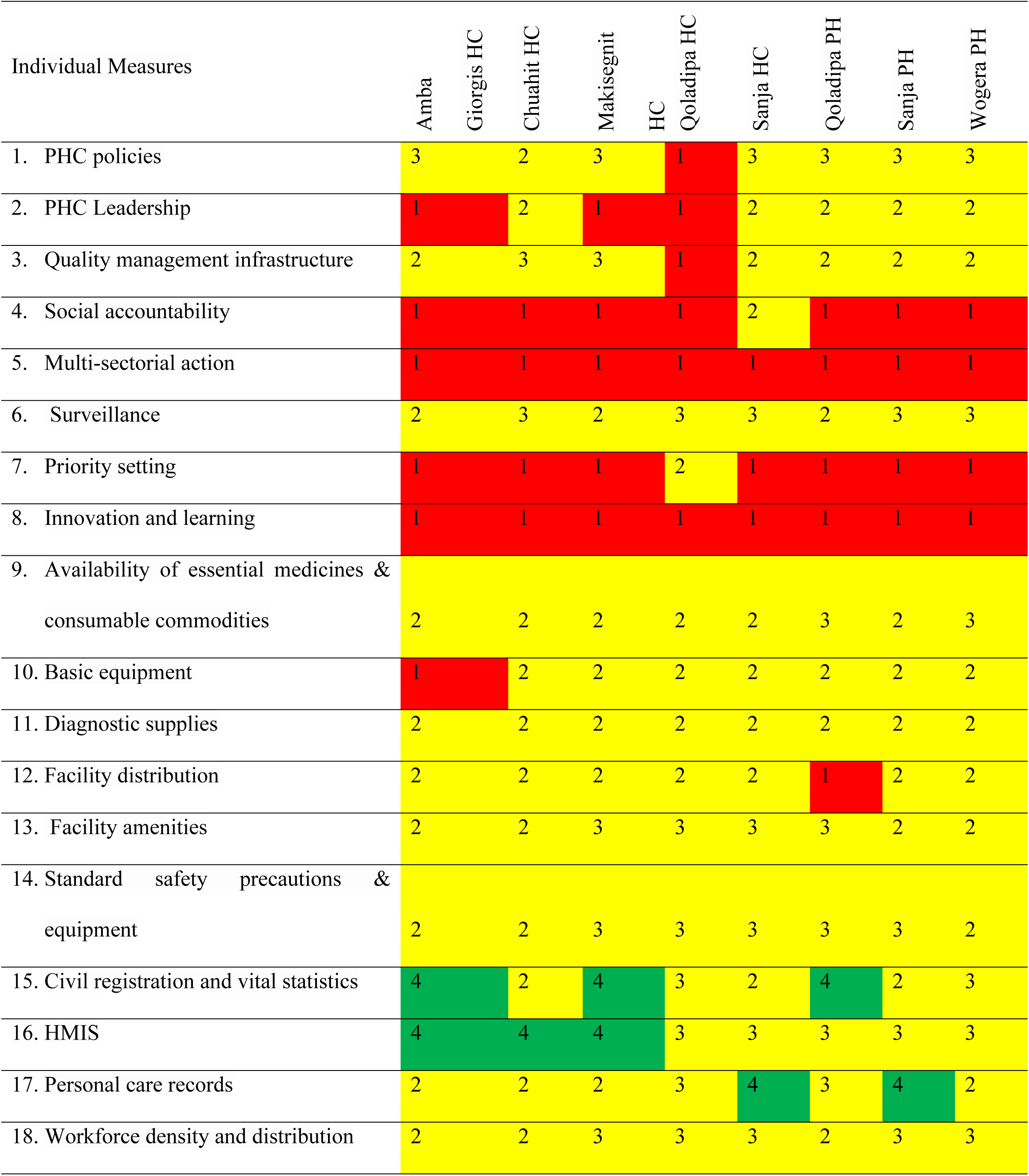
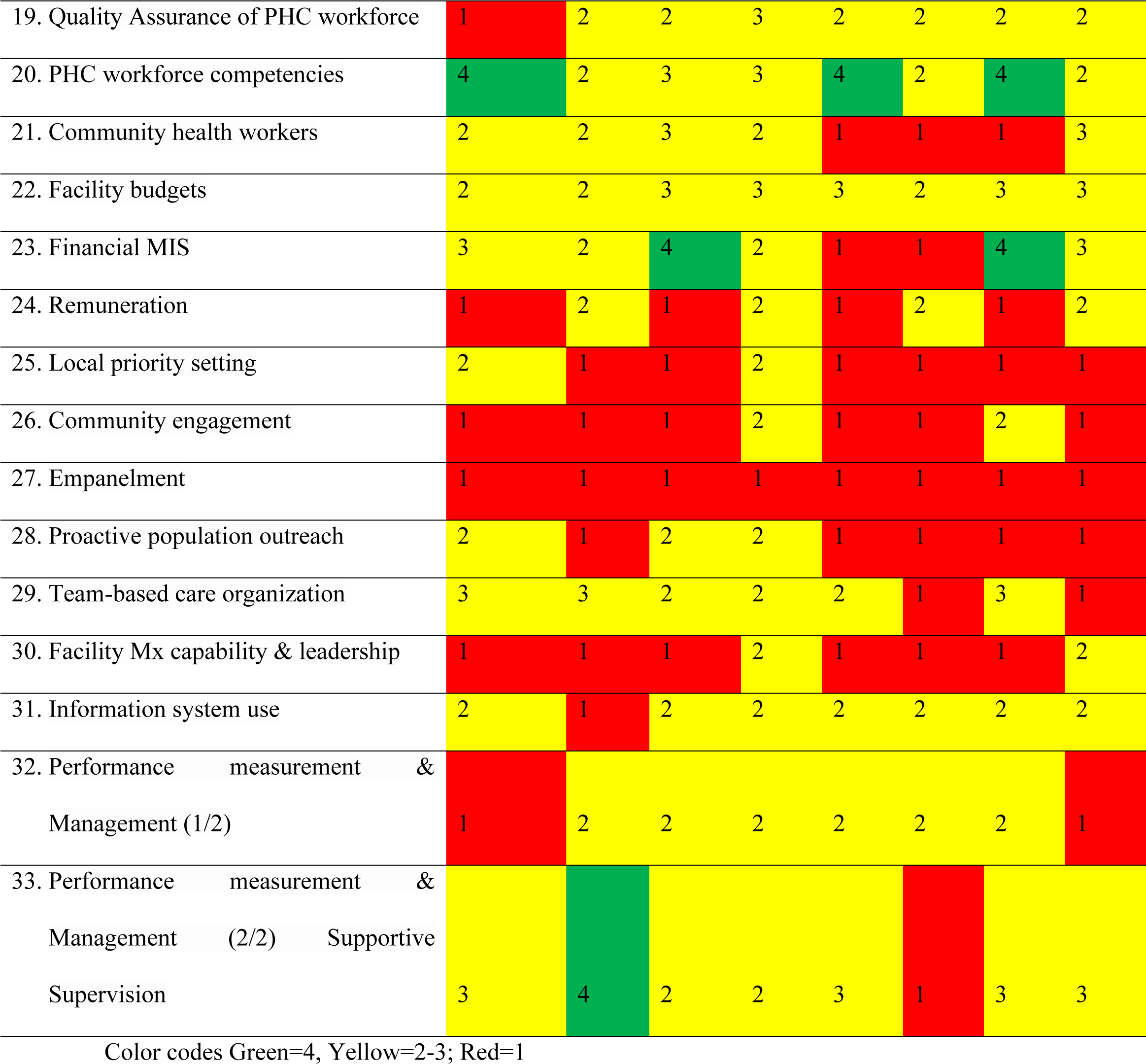
Individual scores out of four for each measure in the Primary Health Care Progression Model by facility in Central Gondar zone, Ethiopia, in 2023.

### The PHC progression model main domain scores at each facility

Governance was better implemented at wogera primary hospital 2.1/4) as compared to other health facilities, the input domain was better implemented at Sanja and wogera primary hospitals and population health and facility management was also better at Qoladba health center (Table 3).

**Table 3:**
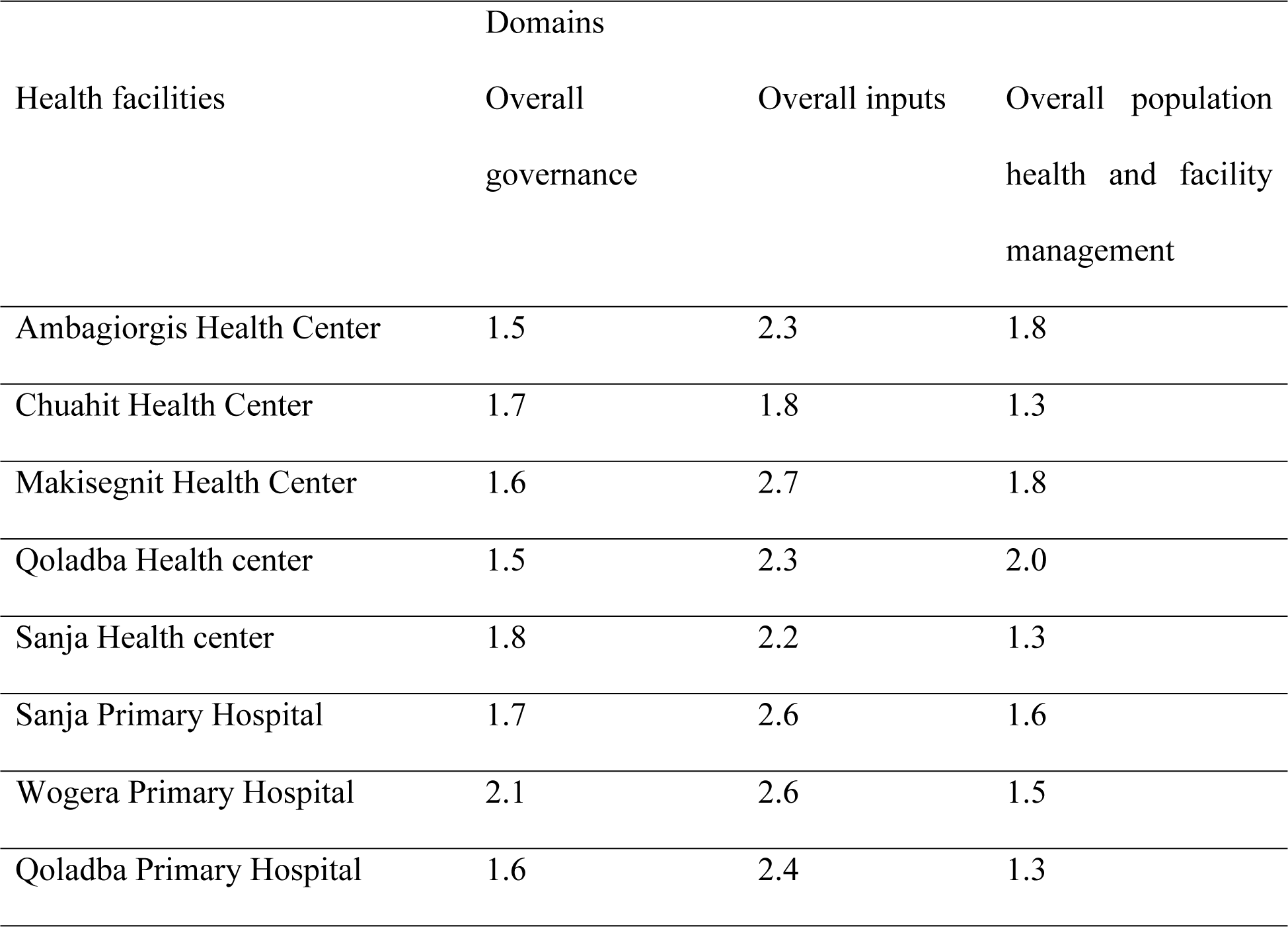
The overall three domains of capacity assessment in Central Gondar, Ethiopia, 2023.

| Health facilities | Domains |  |  |
| --- | --- | --- | --- |
|  | Overall governance | Overall inputs | Overall population health and facility management |
| Ambagiorgis Health Center | 1.5 | 2.3 | 1.8 |
| Chuahit Health Center | 1.7 | 1.8 | 1.3 |
| Makisegnit Health Center | 1.6 | 2.7 | 1.8 |
| Qoladba Health center | 1.5 | 2.3 | 2.0 |
| Sanja Health center | 1.8 | 2.2 | 1.3 |
| Sanja Primary Hospital | 1.7 | 2.6 | 1.6 |
| Wogera Primary Hospital | 2.1 | 2.6 | 1.5 |
| Qoladba Primary Hospital | 1.6 | 2.4 | 1.3 |

### Summarized internal, external and consensus scores with respective average scores

Among primary health care capacity measures, the lowest average internal assessment score was 1 out of 4 (social accountability multi-sectorial action, and innovation and learning whereas the highest score was 3.1 (Surveillance Health Management Information Systems, and personal recorders). The highest average external as well as average consensus score was 3.38 (Health Management Information Systems).

Almost the final internal and external assessment scores were similar and they agreed on the score either consensus or on average score on eight health facilities (Table 4)

**Table 4:**
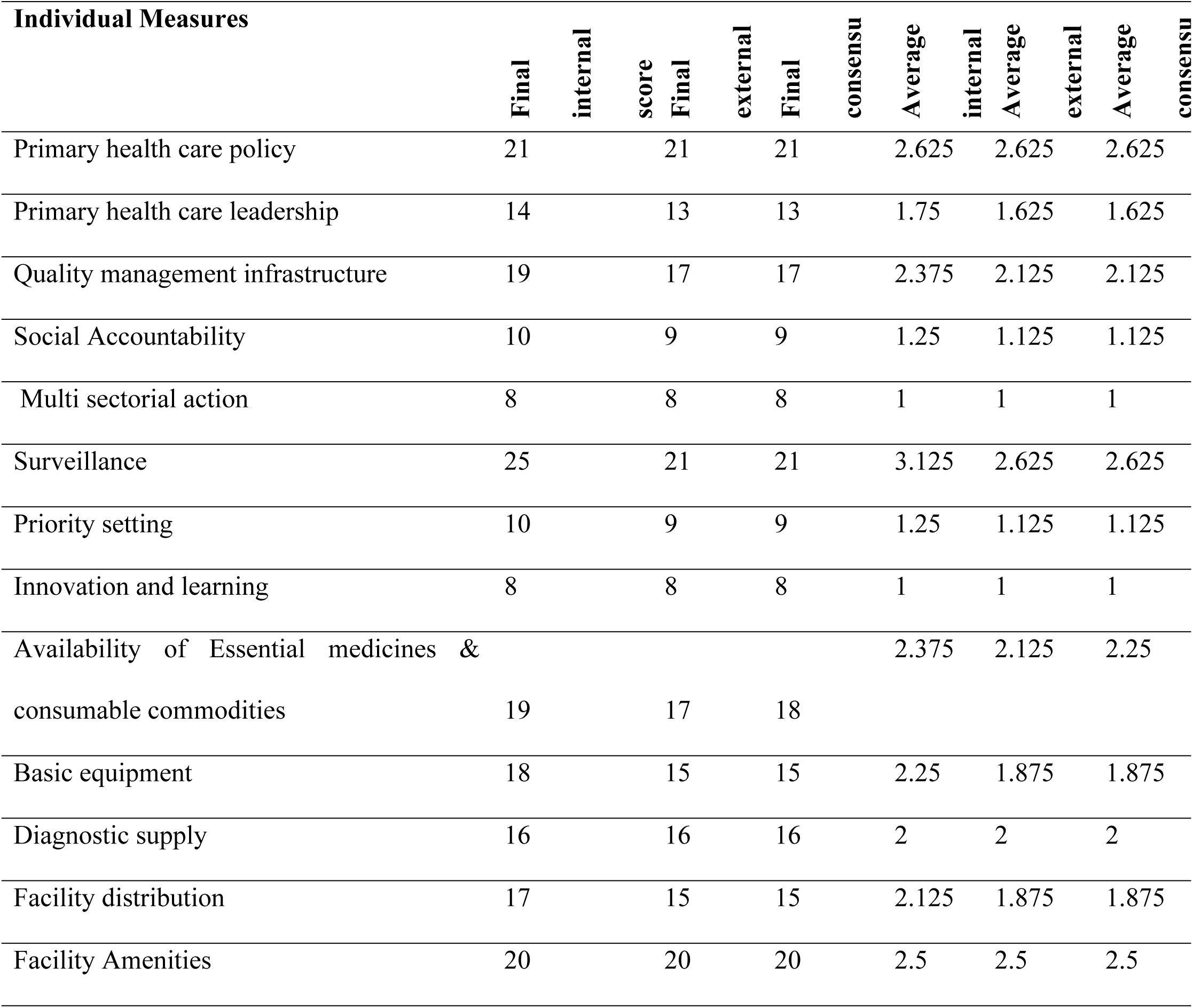
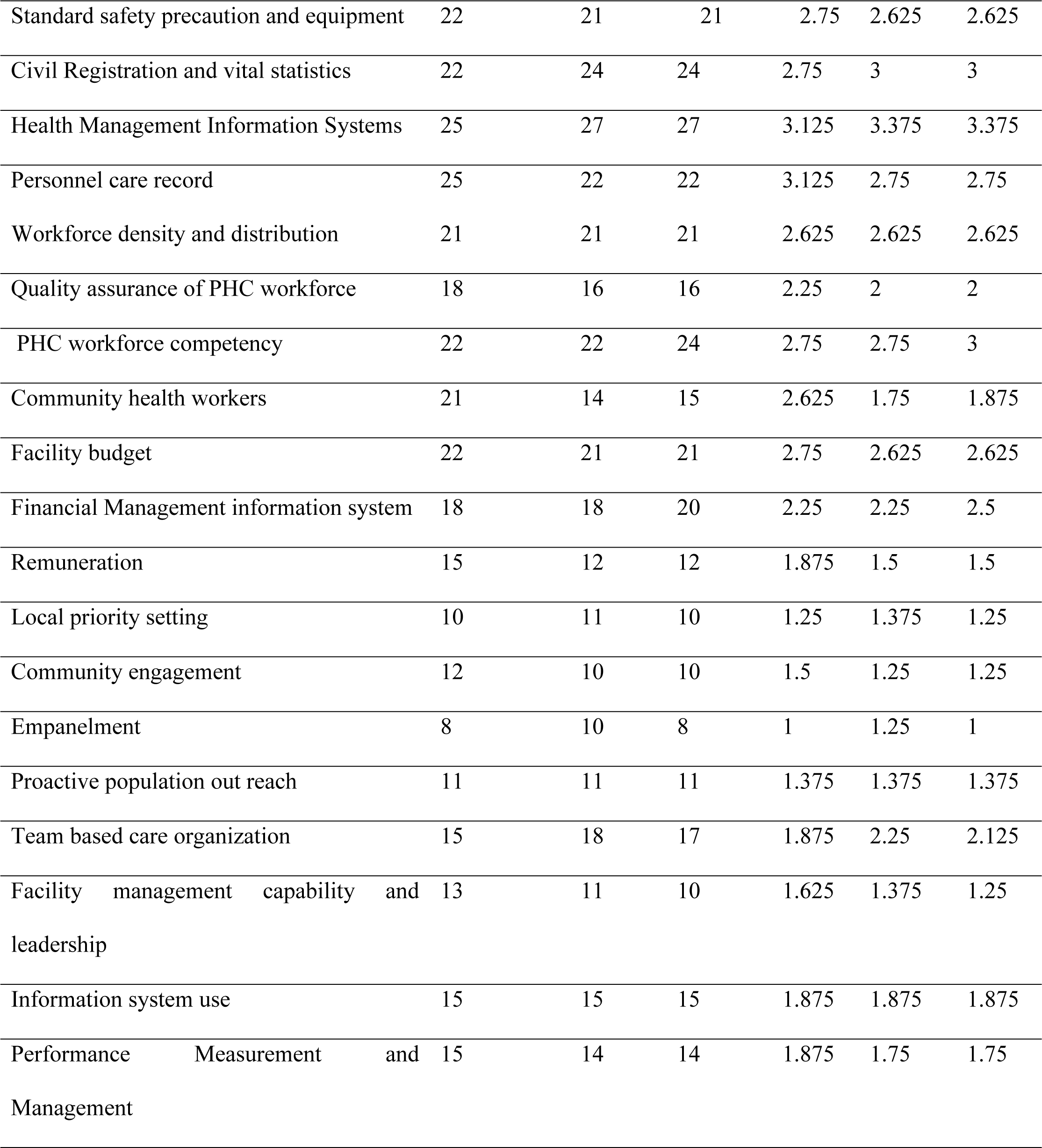
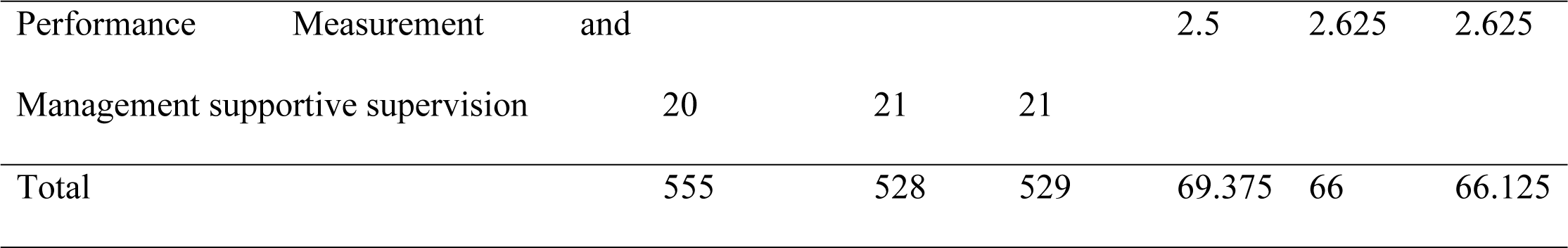
Final internal, external and consensus and average scores of each PHC measure central Gondar Ethiopia, 2023 (Summary of 8 HFs).

### The primary health care progression model subdomains measure scores out of 4 points

The subdomain measures scored by internal and external scorers were: Governance and Leadership, Adjustment to Population Health Needs, Drugs and Supplies, Facility Infrastructure, Information Systems, Workforce, Funds, Population Health Management, and Facility Organization and Management. (Table 5)

**Table 5:**
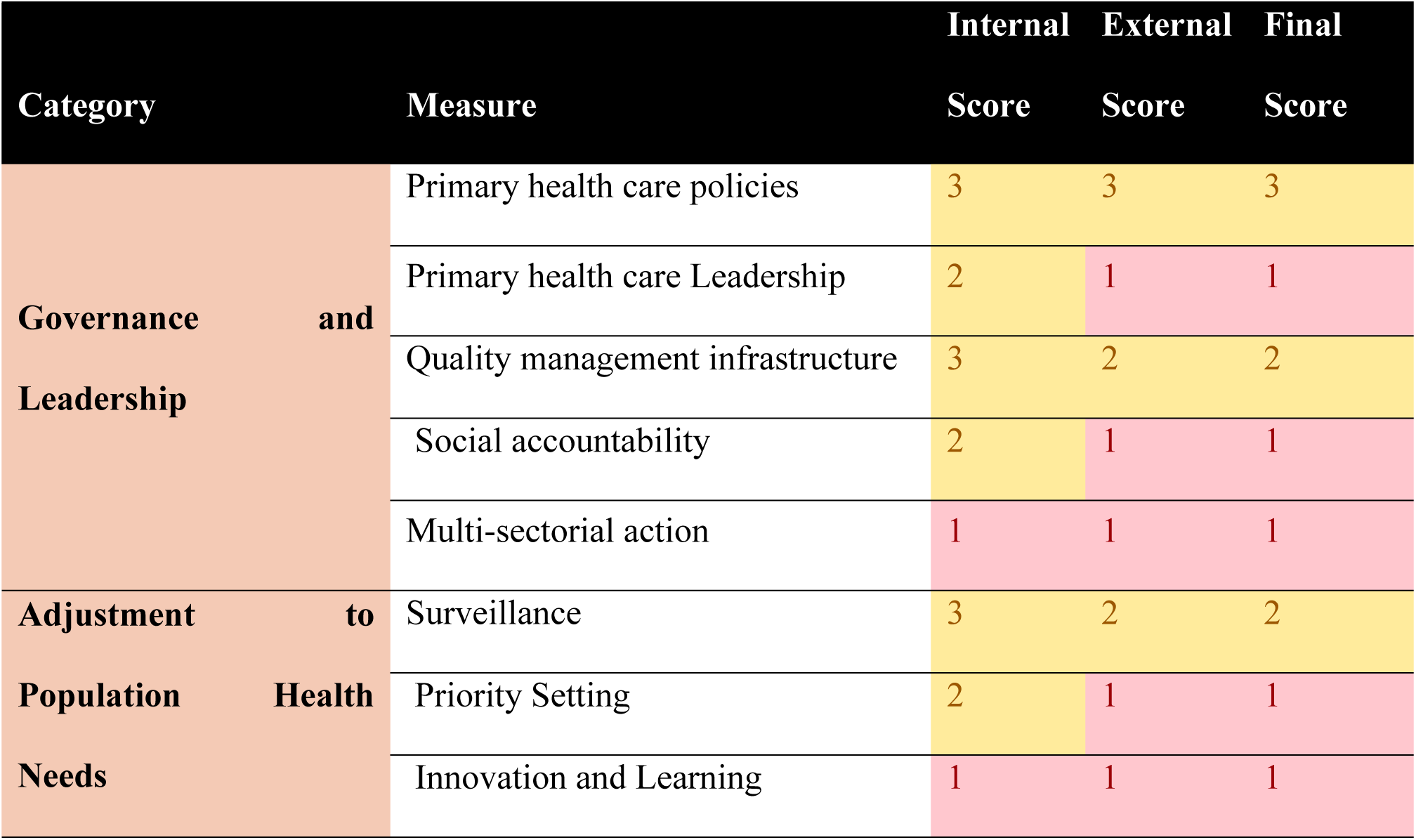
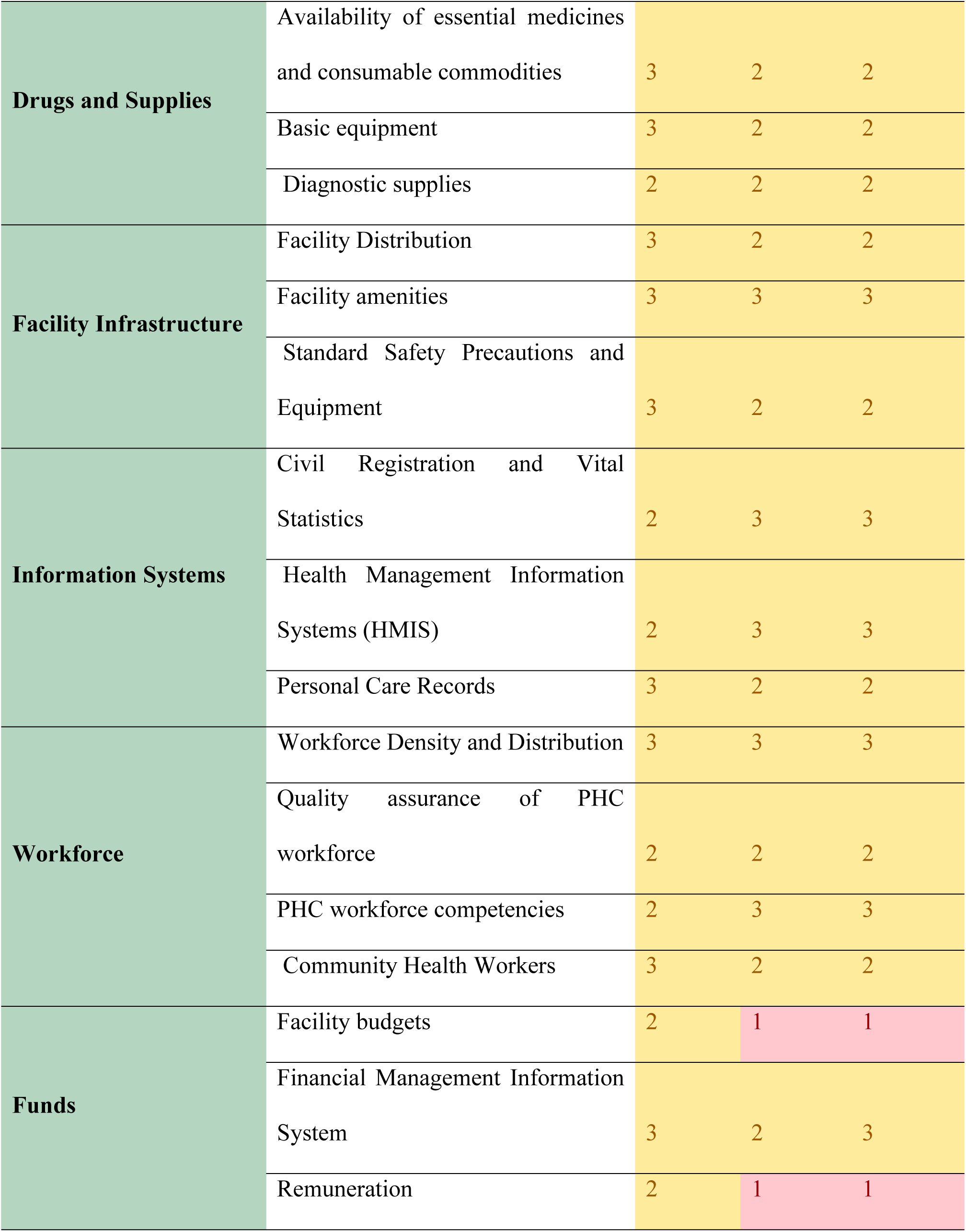
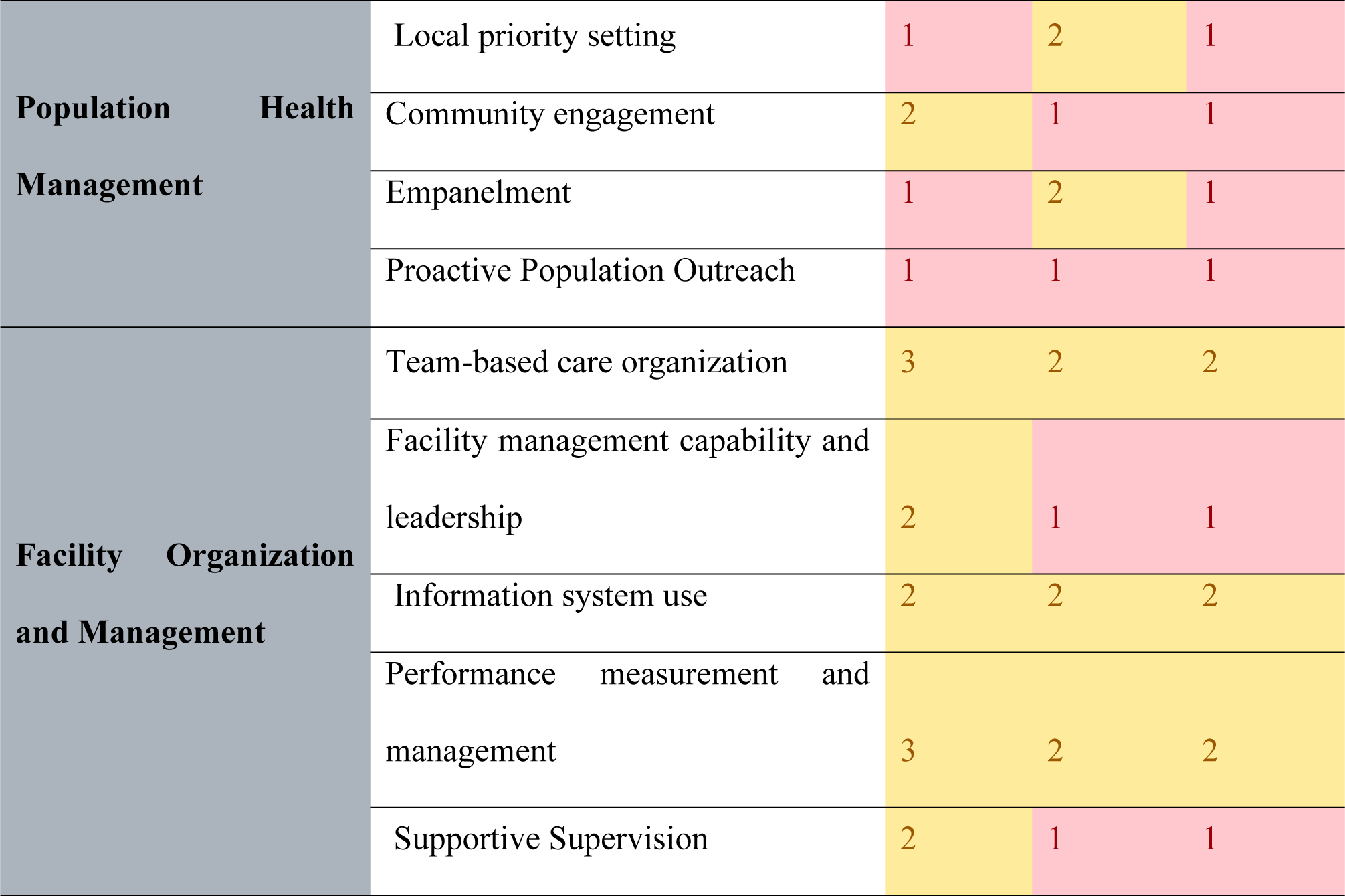
Primary health care progression model subdomain measure scores out of 4 points Central Gondar, Ethiopia, 2023.

## Discussion

This study evaluated the capacity of PHC in health facilities in Central Gondar zone using the PHC progression model. The results indicated a generally low level of PHC capacity across all assessed domains. Significantly, the capacity in the population health and facility management domain was found to be particularly lacking. While the input domain exhibited low scores as well, it was comparatively better than those of the population health management and governance domains. These findings highlight critical areas for improvement in PHC capacity, especially in governance and population health and facility management aspects, which are essential for enhancing overall health system performance.

Utilizing the PHC progression model, the study showed that the overall capacity of health facilities was 1.5 out of 4 in the governance domain, 2.2 out of 4 in the input’s domain, and 1.3 out of 4 in the population health and facility management domain. This finding is low compared to the national level study in Ethiopia, however, the scores of input was similar from the Ministry of Health assessment of capacity[13].

### Governance

The Governance domain of the study achieved an average score of 2.2 out of 4, which is categorized as a medium score according to the Primary Health Care (PHC) progression model. This score is notably lower than the findings of a national level study in Ethiopia, which reported an average score of 2.8 out of 4 [14], and a study conducted in Qatar that scored 3.6 out of 4 [15].

The observed differences in scores may be attributed to variations in the study settings; the previous studies were conducted at a national level focusing on regional offices, while our study was performed at the facility level. The implications of our findings suggest potential issues such as the unavailability of essential policies and strategies, lack of social accountability, inadequate handling of quality documents, violations of rules and regulations, and the absence of quality improvement projects and capacity-building training. These factors may contribute to the suboptimal performance of health facilities [4].

The study assessed eight governance measures in health care including primary health care policy, primary healthcare leadership, social accountability, multi-sectorial action, quality management infrastructure, surveillance, priority setting and innovation and learning. Among these only three domains; primary healthcare leadership, quality management infrastructure and surveillance scored 2 or above out of 4, indicating medium level performance. The remaining domains scored 1 out of 4, highlighting that over 60% of the governance areas are poorly functioning and require urgent attention. The findings suggest that without prompt action from the health care government in central Gondar zone, significant impairments in health service delivery may occur due to limited leader-workers interaction, poor community engagement, inadequate collaboration with public organizations, and in effective participatory decision making[16]. District and zonal health offices must address these gaps urgently and collaborate with health facilities [17]. Evidences indicated that the capacity building training, organizational restructuring, and fostering a positive social environment can enhance leadership capacities in health facilities[18].

### Inputs

The input dimension of PHC capacity received an overall score of 2.2 out of 4, indicating a medium level performance. Within this dimension, the lowest score was 1.7, attributed to funding, while the highest score was 2.7, related to information system.

In this study, the financial aspect of the domain received the lowest score among all subdomains. Suggest that delivering quality health services is challenging due to the dynamic nature of healthcare, which requires constant adaptation to emerging patient needs, new technologies, and changing environments. Evidences indicate that robust healthcare budget is crucial for leaders to effectively plan for the future and prioritize care across various departments and programs, ultimately enabling health care organizations to provide more efficient and effective patient care [19]. Healthcare budgeting plays a vital role in decision making with in health system and organizations. An effective budgeting process enables health care leaders to align daily operations with financial targets, prioritize key areas in line with strategic goals and manage capital expenditures and cash flow efficiently. Additionally, it provides a clear understanding of funding allocation for individual projects, initiatives and clinical departments, while also helping to minimize purchasing errors[20].

The study identified several budget related challenges faced by health centers and hospitals in central Gondar zone. These challenges include unpredictable economic events such as the COVID 19 pandemic and conflicts which render static budgets ineffective. Other issues include changes or delay in reimbursement and payment models, labor shortage, rising costs and supply chain disruptions, particularly shortage of personal protective equipment (PPE) and prescribed drugs. In addition, external factors such as fluctuation in patient volume and inflation also significantly impact health care budget[21].

In the input domain, the availability of essential medicines and consumable commodities received score of 2 and 3 out of 4 across all facilities, categorizing them at the “yellow level.” This finding aligns with a systematic review conducted in low and middle income countries[22]. Stock-outs of medical consumables adversely affect health outcomes by delaying effective service delivery and discouraging patients from seeking care. The study also revealed low availability of fully functioning basic equipment, with Ambagiorgis health center scoring “red” (1/4) and other facilities scoring “yellow” (2/4). This outcome is consistent with a study from Jimma, Ethiopia, which found that one-third of basic equipment was unavailable [23].

The civil registration and vital statistics measure achieved a "green level" score (4/4) at Ambagiorgis, Makisegnit, and Qoladba health centers, indicating a high level of performance in these facilities. In contrast, other health facilities only reached a "yellow level," suggesting room for improvement. This disparity may be linked to the government’s prioritization of civil registration and vital statistics, along with effective collaboration between health facilities and government administration offices in Ethiopia.

Additionally, the Management and Health Information Systems (MHIS) measure demonstrated strong performance, achieving a "green level" score (4/4) at three health facilities and a "yellow level" (3/4) at five others. This performance is consistent with findings from a study conducted in public hospitals across Ethiopia[24], although it surpasses results from a study in the Oromia zone. The variation in results may stem from the different study tools employed; our study utilized indicators from the PHC Progression Model, while the previous study relied on indicators from national HMIS guidelines[25].

### Population health and facility management

The population health and facility management domain of primary health care (PHC) received a low overall score of 1.3 out of 4. Nine measures were evaluated by both internal and external assessors, with the final scores reached by consensus. Only three measures—team-based care, information system use, and performance measurement—scored medium based on primary health care progression models. In contrast, several critical measures, including local priority setting, community engagement, proactive population outreach, empanelment, and supportive supervision, were poorly implemented in the study area, highlighting significant gaps in health system priorities[4].

The findings indicate that local community health issues are often overlooked, leading to decision-making and resource allocation being influenced more by historical and political factors than by technical knowledge or an understanding of current health needs [26]. Community engagement, recognized by the World Health Organization as a key measure of primary health care capacity, is highlighted as a cost-effective strategy for promoting health and preventing illness. Although there are no direct comparative studies, various preliminary findings and reports from international organizations emphasize the importance of community engagement in local health governance. Active participation of residents not only enhances democratic governance but also increases their support for health initiatives. Moreover, community engagement fosters strong, cohesive communities where residents feel connected and supported. This sense of belonging can reduce social isolation, increase cooperation, and promote a spirit of mutual aid and collective responsibility. It also enables local governments to create more sustainable and inclusive policies, as engaged communities are more likely to support and participate in initiatives related to environmental conservation, economic development, and social well-being.

During crises, communities that are actively engaged are more capable of responding effectively to challenges such as natural disasters, economic difficulties, or public health emergencies. Additionally, community engagement fosters a sense of ownership among residents in local governance, resulting in increased civic pride, higher volunteerism, and a more dynamic citizenry [27]. However, our findings indicate that the level of implementation in the central Gondar zone is low (1 out of 4). Specifically, there have been no community forums, campaigns, or activities aimed at building trust between the community and the health system. These shortcomings may be attributed to issues of insecurity and the adverse effects of the COVID-19 pandemic.

## Data Availability

All relevant data are within the manuscript and its Supporting Information files.

## Acknowledgement

We authors would like to thank to the University of Gondar, internal and external assessors for devoting their time and commitment, the study participants for providing information and their patience

## Authors’ contribution

**Conceptualization:** Chalie Tadie Tsehay, Tsegaye G. Haile, Nigusu Worku, Asmamaw Atnafu.

**Data curation**: Chalie Tadie Tsehay, Nigusu Worku, Wubshet Debebe Negash

**Formal analysis:** Chalie Tadie Tsehay, Nigusu Worku

**Methodology:** Chalie Tadie Tsehay, Tsegaye G.Haile, Endalkachew Delie,Wubesht Debebe Negash,Andualem Yalew Aschalew, Ayal Debie, Samrawit Mihret Fetene,Adana Kebede, Asmamaw Atnafu

**Validation:** Chalie Tadie Tsehay, Tsegaye G.Haile, Endalkachew Delie,Wubesht Debebe,Nigusu Worku, Asmamaw Atnafu

**Visualization:** Chalie Tadie Tsehay, Tsegaye G.Haile, Endalkachew Delie,Wubesht Debebe Negash,Andualem Yalew Aschalew, Ayal Debie, Samrawit Mihret Fetene,Adana Kebede, Asmamaw Atnafu

**Writing original draft:** Chalie Tadie Tsehay, Nigusu Worku

